# Unhealthy Behaviours and Parkinson’s Disease: A Mendelian Randomisation Study

**DOI:** 10.1101/2020.03.25.20039230

**Authors:** Karl Heilbron, Melanie P. Jensen, Sara Bandres-Ciga, Pierre Fontanillas, The 23andMe Research Team, Cornelis Blauwendraat, Mike A. Nalls, Andrew B. Singleton, George Davey Smith, Paul Cannon, Alastair Noyce

## Abstract

**Objective:** Tobacco smoking, alcohol intake, and high BMI have been identified in observational studies as potentially protective factors against developing Parkinson’s disease (PD). Because of the possibility of residual confounding and reverse causation, it is unclear whether such epidemiological associations are causal. Mendelian randomisation (MR) uses genetic variants to explore causal effects of exposures on outcomes; minimising these sources of bias. Using MR, this study sought to determine the causal relationship between tobacco smoking, alcohol intake, and high BMI, and the risk of PD.

**Methods:** We performed genome-wide association studies to identify single nucleotide polymorphisms associated with the exposures. MR analysis of the relationship between each exposure and PD was undertaken using a split-sample design. The inverse variance weighted (IVW) method was used to combine SNP-specific effect estimates.

**Results:** Ever-smoking causally reduced risk of PD (OR 0.955; 95% confidence interval [CI] 0.921-0.991; p=0.013). An increase in daily alcohol intake causally increased risk of PD (OR 1.125, 95% CI 1.025-1.235; p=0.013) and a 1 kg/m^2^ BMI causally reduced risk of PD (OR 0.988, 95% CI 0.979-0.997; p=0.008). Sensitivity analyses did not suggest bias from horizontal pleiotropy or invalid instruments.

**Interpretation:** Using split-sample MR in over 2.4 million participants, we observed a protective effect of smoking on risk of PD, warranting the prioritisation of related therapeutic targets, such as nicotinic agonists, in prevention trials. In contrast to observational data, alcohol consumption causally increased risk of PD. Higher BMI had a protective effect on PD, but the effect was small.

## Introduction

Parkinson’s disease (PD) is the second most common neurodegenerative disorder, and the number of people living with PD is set to double from 6.9 million in 2015 to 14.2 million in 2040.^1^ The mechanisms driving the cardinal pathology of PD – alpha-synucleinopathy and dopaminergic nigrostriatal neurodegeneration – remain unclear. Accordingly, existing therapeutic approaches are symptomatic and have no discernible impact on disease process. Observational studies have identified putative modifiable risk factors for PD but their causal effect on aetio-pathogenesis, hence their value as therapeutic targets for neuroprotection, has not been elucidated.

Tobacco use and alcohol intake are notable as potentially protective factors against being diagnosed with PD, but have detrimental effects on other health outcomes. Observational studies suggest that ever-smokers are 40% less likely to develop PD than never-smokers.^2^ The purported association is plausible given the observed dose-response relationship with pack-years, and replication of the protective effect for oral smokeless tobacco and dietary nicotine.^2-4^ Similar associations have been reported with alcohol intake: high versus low (smoking-adjusted) alcohol intake was associated with a 22% lower risk of PD in a meta-analysis involving 670,550 subjects.^5^ A recent large-scale, case-control study supported the negative observational associations with both smoking and alcohol.^6^

The role of body mass index (BMI) in risk of PD is less clear. Low BMI is apparent in patients with PD in case-control studies, however a meta-analysis of 10 prospective studies found no association between premorbid BMI and PD risk.^7, 8^ Conversely, a recent Mendelian randomisation (MR) study pointed towards a protective effect of higher BMI, which was not clearly due to survival bias.^9^

It is unclear whether such epidemiological associations are truly causal. Risk-averse individuals may be both less likely to engage in unhealthy behaviours and more likely to develop PD, leading to unmeasured confounding and/or reverse causation.^10^ Reverse causation is plausible given the long prodromal phase of PD.^11^ Indeed, ease of smoking cessation and weight loss may be prodromal features of PD.^12, 13^ Such factors may explain why randomised controlled trials (RCTs) and crossover trials have failed to demonstrate any disease-modifying or symptomatic benefit of nicotine on PD.^14-18^ However, these trials relate to disease progression, rather than disease risk, and the events driving disease progression may be different to those initiating it.^19^

Using MR, this study sought evidence for a causal relationship between PD risk (outcome) and three unhealthy behaviours (exposures): tobacco smoking, alcohol intake, and high BMI. In MR, genetic variants associated with the exposure of interest are used as instrumental variables to estimate the effect of the exposure on the outcome.^20, 21^ The random allocation of genetic variants from parent to offspring means that alleles are generally unrelated to confounding factors (measured and unmeasured) which distort inferences from observational data.^22^ Moreover, the germline genotype cannot be modified by the disease process, minimising bias from reverse causality.^22^ MR also addresses limitations of RCTs, both ethical (when participants may be assigned to a harmful exposure, *e.g*. smoking) and pragmatic (when adherence cannot be guaranteed, *e.g*. alcohol intake, BMI modulation).^23^

## Methods

### Participants

Participants were customers of 23andMe, Inc., a personal genomics company. Study protocols were approved by an external AAHRPP-accredited institutional review board and conducted in accordance with the Declaration of Helsinki principles. Participants gave informed consent to participate. Data was collected between November 10th, 2007 and January 1st, 2018.

### Defining phenotypic traits

#### Parkinson’s disease status

PD cases were drawn from the 23andMe participants who self-reported a diagnosis of PD. For a fuller description of recruitment see Do *et al*.^*24*^ A high level of agreement between patient-reported PD diagnosis and neurologist assessment has previously been demonstrated.^25^ Controls were drawn from the 23andMe participants who self-reported never having been diagnosed with PD. 19,924 PD cases and 2,413,087 controls were included in the analysis.

#### Unhealthy exposures of interest

Participants self-reported their tobacco smoking habits with response categories “ever-smoker (>100 cigarettes smoked in lifetime)”, or “never-smoker (<100 cigarettes smoked in lifetime)”. Participants self-reported the number of alcohol measures consumed per day over the past two weeks, where 1 measure corresponds to 12 oz. of beer, 5 oz. of wine, or 1.5 oz of spirits. Possible response options were grouped as follows: 0 measures, 0-1 measure, 1 measure, 2 measures, 3 measures, 4 measures, 5 or more measures. Participants self-reported their mass (in kilograms) and height (in metres) squared, from which BMI was calculated.

### Genome-wide association studies

We performed new genome-wide association studies for each phenotypic exposure using unrelated individuals. We selected unrelated individuals using a segmental identity-by-descent estimation algorithm.^26^ Individuals were defined as related if they shared 700 cM identity-by-descent, including regions where the two individuals share either one or both genomic segments identical-by-descent. This level of relatedness (roughly 20% of the genome) corresponds approximately to the minimal expected sharing between first cousins in an outbred population. Ancestry composition was performed as previously reported.^27^ Inclusion was restricted to individuals of predominantly European ancestry to minimise confounding by ancestry.^28^

DNA extraction and genotyping were performed on saliva samples by National Genetics Institute. Samples were genotyped on one of five Illumina-based genotyping platforms. The v1 and v2 platforms were variants of the Illumina HumanHap550+ BeadChip, including about 25,000 custom SNPs selected by 23andMe, with a total of about 560,000 SNPs. The v3 platform was based on the Illumina OmniExpress+ BeadChip, with custom content to improve the overlap with our v2 array, with a total of about 950,000 SNPs. The v4 platform was a fully customized array, including a lower redundancy subset of v2 and v3 SNPs with additional coverage of lower-frequency coding variation, and about 570,000 SNPs. The v5 platform is an Illumina Infinium Global Screening Array (∼640,000 SNPs) supplemented with ∼50,000 SNPs of custom content. Samples had minimum call rates of 98.5%.

We phased participant data using either an internally-developed tool, Finch (V1-V4 genotyping arrays) or Eagle2 (V5 genotyping array).^29^ Finch implements the Beagle haplotype graph-based phasing algorithm, modified to separate the haplotype graph construction and phasing steps.^30^ It extends the Beagle model to accommodate genotyping error and recombination, to handle cases where there are no consistent paths through the haplotype graph for the individual being phased. We constructed haplotype graphs for European and non-European samples on each 23andMe genotyping platform from a representative sample of genotyped individuals, and then performed out-of-sample phasing of all genotyped individuals against the appropriate graph. For the X chromosome, we built separate haplotype graphs for the non-pseudoautosomal region and each pseudoautosomal region, and these regions were phased separately.

#### Imputation

Imputation panels created by combining multiple smaller panels have been shown to give better imputation performance than the individual constituent panels alone.^31^ To that end, we combined the May 2015 release of the 1000 Genomes Phase 3 haplotypes with the UK10K imputation reference panel to create a single unified imputation reference panel.^32, 33^ To do this, multiallelic sites with N alternate alleles were split into N separate biallelic sites. We then removed any site whose minor allele appeared in only one sample. For each chromosome, we used Minimac3 to impute the reference panels against each other, reporting the best-guess genotype at each site.^34^ This gave us calls for all samples over a single unified set of variants. We then joined these together to get, for each chromosome, a single VCF with phased calls at every site for 6,285 samples.

In preparation for imputation we split each chromosome of the reference panel into chunks of no more than 300,000 variants, with overlaps of 10,000 variants on each side. We used a single batch of 10,000 individuals to estimate Minimac3 imputation model parameters for each chunk.^34^ We imputed phased participant data against the chunked merged reference panel using Minimac3, treating males as homozygous pseudo-diploids for the non-pseudoautosomal region. Throughout, we treated structural variants and small indels the same as SNPs.

#### Association tests

We computed association test results by regression assuming additive allelic effects (logistic regression for case-control exposures, linear regression for quantitative exposures). We included covariates for age, sex, the top five genetic principal components to account for residual population structure, and indicators for genotype platforms to account for genotype batch effects. The association test p-value we report was computed using a likelihood ratio test. For tests using imputed data, we use the imputed dosages rather than best-guess genotypes. For the X chromosome, male genotypes were coded as if they were homozygous diploid for the observed allele.

#### Principal component analysis

We performed the genetic principal components analysis using ∼65,000 high-quality genotyped variants present in all five genotyping platforms and a random sample of one million research participants with predominantly European ancestry. Principal component scores for participants not included in the analysis were obtained by projection, combining the eigenvectors of the analysis and the SNP weights.

#### Quality control of genotyped GWAS results

We excluded SNPs that: 1) had a call rate <90%, 2) had a Hardy-Weinberg *P*<10^−20^ in people with predominantly European ancestry, 3) were only genotyped on the V1 and/or V2 platforms, 4) were found on the mitochondrial chromosome or the Y chromosome, 5) failed a test for parent-offspring transmission (specifically, we regressed the child’s allele count against the mean parental allele count and excluded SNPs with fitted β<0.6 and P<10^−20^ for a test of β<1), 6) had an association with genotype date (P<10^−50^ by ANOVA of SNP genotypes against a factor dividing genotyping date into 20 roughly equal-sized buckets), 7) had a large sex effect (ANOVA of SNP genotypes, r^2^>0.1), or 8) had probes matching multiple genomic positions in the reference genome.

#### Quality control of imputed GWAS results

We excluded SNPs with imputed r^2^<0.3, as well as SNPs that had strong evidence of a platform batch effect. For each SNP we identified the largest subset of the data passing other quality control criteria based on their original genotyping platform – either v2+v3+v4+v5, v4+v5, v4, or v5 only – and computed association test results for the largest passing set. The batch effect test is an F test from an ANOVA of the SNP dosages against a factor representing the V4 or V5 platform; we excluded results with P<10^−50^.

#### Additional quality control of GWAS results

Across both genotyped and imputed GWAS results, we excluded SNPs that had sample size of less than 20% of the total GWAS sample size. We also removed SNPs that did not converge during logistic regression, as identified by abs(effect)>10 or stderr>10 on the log odds scale. We removed SNPs with MAF < 0.1% from linear regressions because these SNPs are sensitive to violations of the regression assumption of normally distributed residuals. If SNPs were both genotyped and imputed, and they passed QC for both, we used results from the imputed analysis. After quality control, we had analysed 904,040 genotyped SNPs and 25,208,208 imputed SNPs.

### Instrument construction

SNPs associated with each of the exposures at the genome-wide significance level (p<5e^-8^) were included as instrumental variables. We excluded SNPs with a minor allele frequency <3% and SNPs in the *HLA* locus (hg19, chromosome 6, 26.0Mbp - 33.7Mbp). Instrument strength was assessed using the mean F statistic, as calculated by the system_metrics function in the TwoSampleMR R package.^35^

### Split-sample MR analysis

We performed MR analysis of the relationship between the exposures of interest and PD using a split-sample design, in accordance with published methods.^22, 36^ Individuals from the 23andMe cohort were randomly allocated into two evenly sized groups (Table 1). The instrument-exposure association was measured in the first group (fold 1), and the instrument-outcome association was measured in the second group (fold 2), and MR analyses undertaken. We then repeated the MR analyses, but using fold 1 for the outcome and fold 2 for the exposure. For each MR method, this resulted in two independent MR estimates, which were combined using an inverse variance weighted (IVW) fixed-effects meta-analysis.

**Table 1.** Table legend: Participant numbers in fold 1 and fold 2.

|  | **Exposure variables** |  |  | **Outcome variable** |
| --- | --- | --- | --- | --- |
|  | Smoking | Alcohol | BMI | Parkinson's |
| **Fold 1** | Ever-smokers: 533,995 | 1,203,903 | 1,268,201 | Cases: 9,852 |
|  | Never-smokers: 728,610 |  |  | Controls: 1,206,650 |
| **Fold 2** | Ever-smokers: 533,704 | 1,204,410 | 1,267,856 | Cases: 10,072 |
|  | Never-smokers: 728,847 |  |  | Controls: 1,206,437 |

The effect of an exposure on PD was calculated for each SNP using the Wald ratio method.^37^ In the IVW analysis we performed a linear regression constrained through the origin of the variant-exposure and the variant-outcome associations for each instrument, weighted by their inverse variance (Table 2).

**Table 2.**
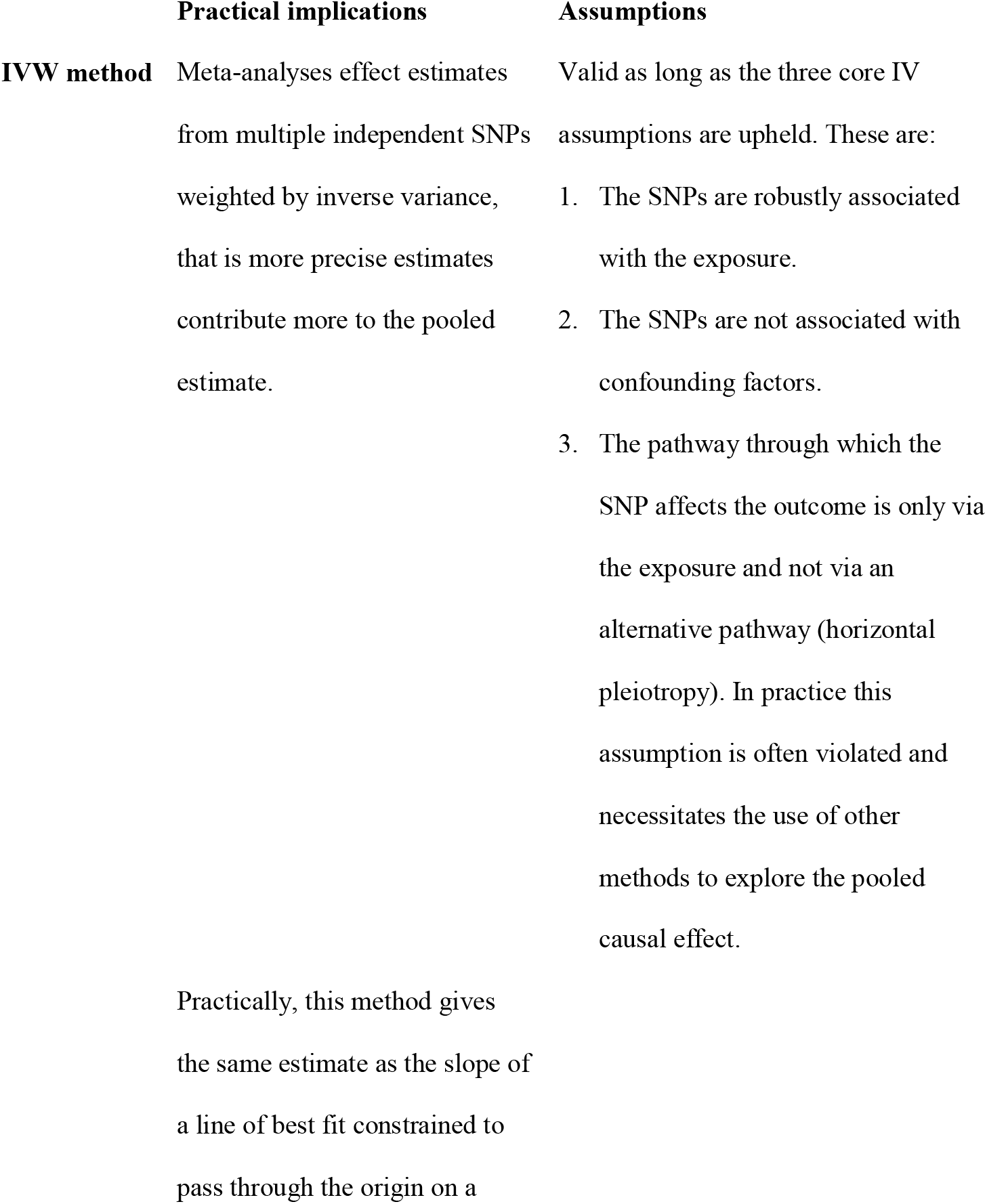
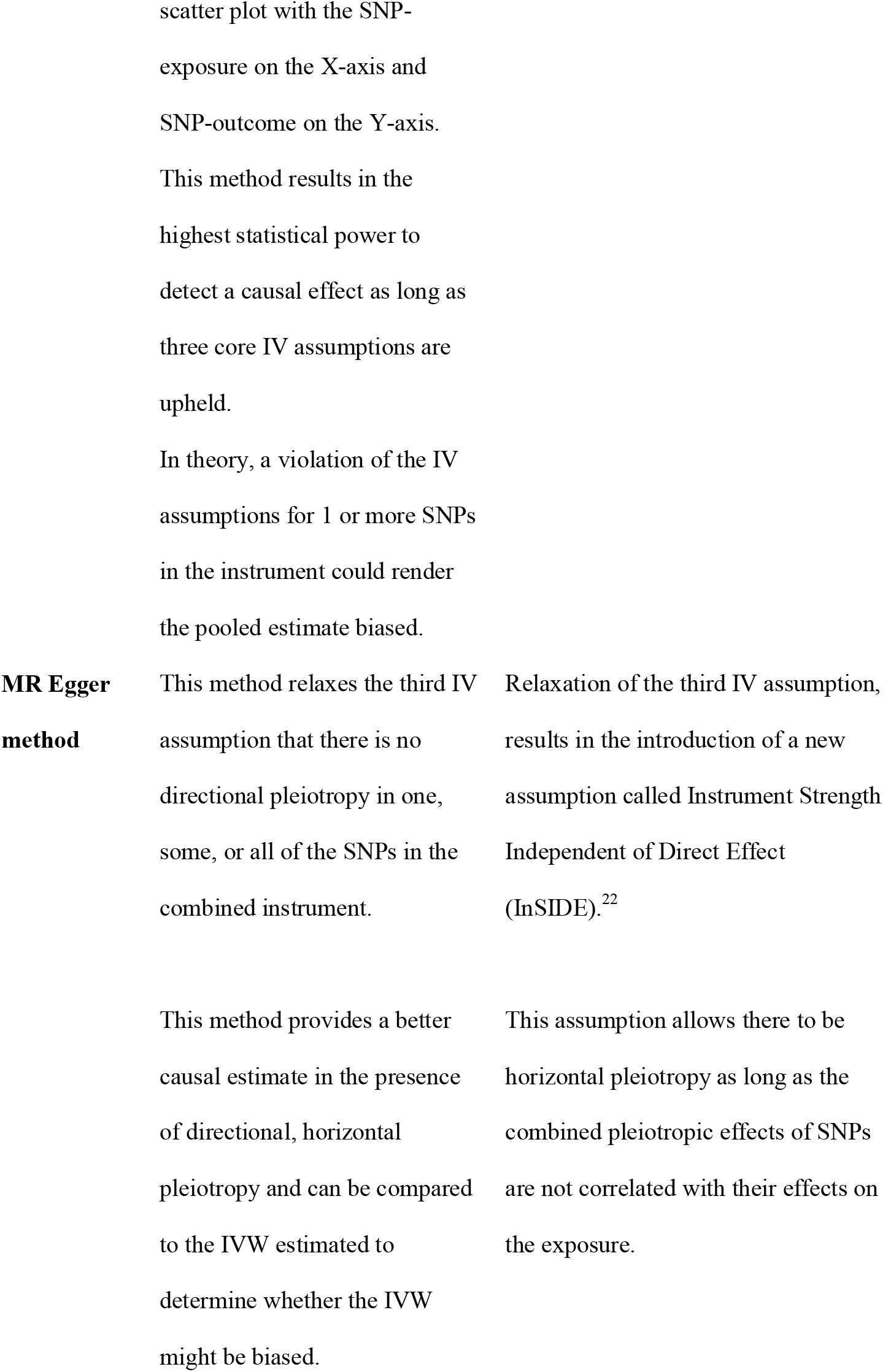
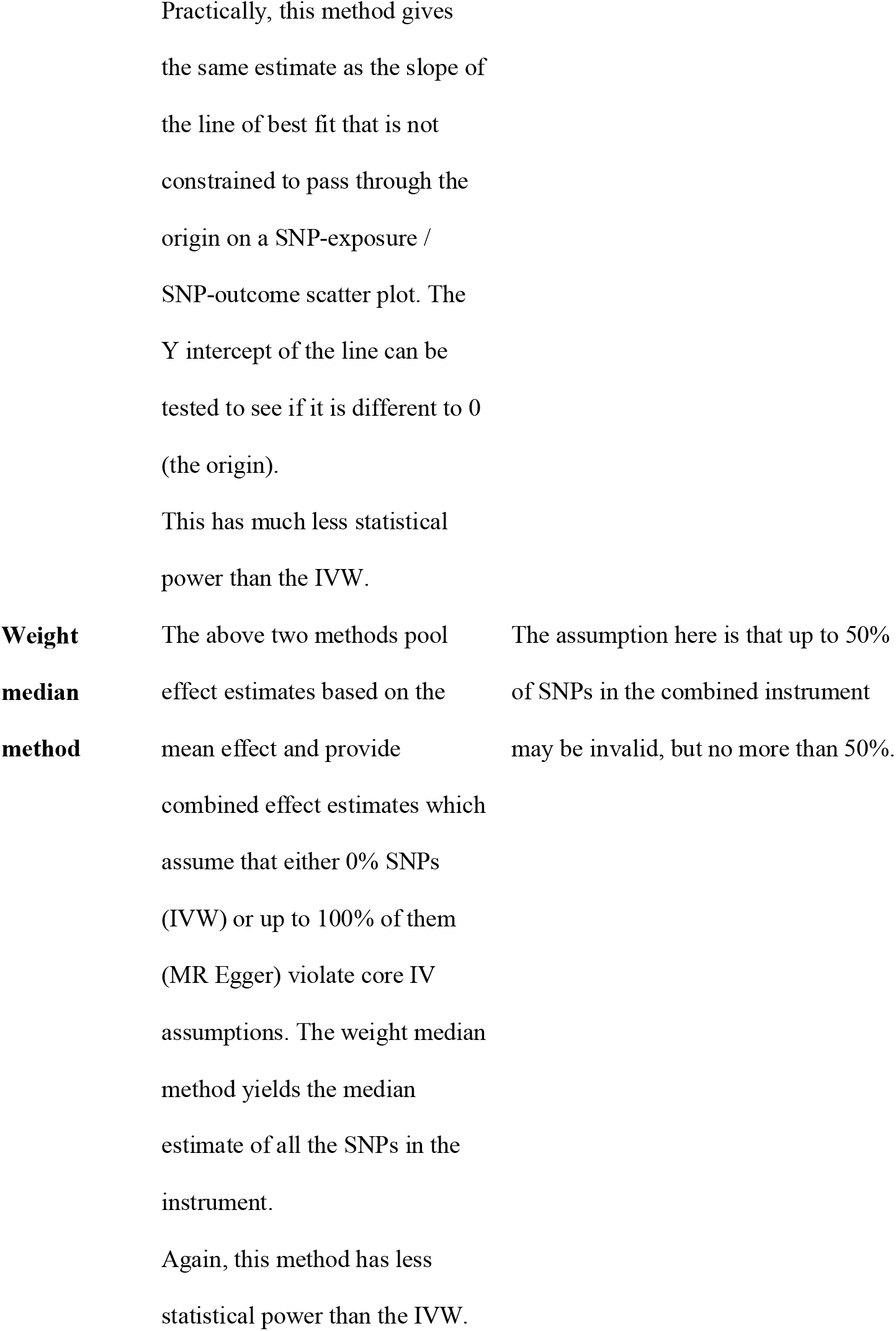
Table legend: Further detail on the characteristics of MR methods used to pool effect estimates. Abbreviations: IVW = inverse variance weight; SNP = single nucleotide polymorphism; IV = instrumental variable.

We used three methods to assess the impact of bias on IVW estimates.^35^ First, heterogeneity in Wald ratio estimates was assessed using the Cochran’s Q test and I^2^ index.^38^ Heterogeneity can indicate the possibility of bias due to horizontal pleiotropy, whereby variants influence the outcome via pathways unrelated to the exposure. Individual variant contributions to Cochran’s Q statistic was calculated, and variants were excluded in the heterogeneity filtering analysis if their contribution surpassed the Bonferroni-corrected 99.8^th^ percentile of the χ^2^(1 df) distribution.

Second, we performed MR-Egger analysis to assess the magnitude of bias in the IVW estimate occurring due to directional horizontal pleiotropy (Table 2). A hypothesis test can be used to determine whether the intercept is different from zero and this indicates a net horizontal pleiotropic effect, but is generally underpowered. In the absence of unbalanced pleiotropy, a funnel plot of the individual variant effects plotted against the inverse of their standard error will be symmetrically distributed around the point estimate.

Third, we used the weighted median method, which reports a valid MR estimate under conditions where up to 50% of the weight of the instruments are invalid (Table 2).^39^ The estimate is obtained by calculating the Wald ratio for each SNP and selecting the median value after weighting by inverse variance. Confidence intervals for the weighted median estimates are calculated through bootstrapping.

To minimize misallocation of variants to the exposure rather than outcome group, we applied Steiger filtering to all analyses to remove genetic variants that had a stronger correlation with the outcome than with the exposure.^40^

### Replication in the International Parkinson’s Disease Genomics Consortium (IPDGC) dataset

Summary statistics from the largest published PD GWAS meta-analysis were used as the outcome data for replication purposes after excluding data from 23andMe (the “IPDGC dataset”).^41^ The outcome summary statistics used for this analysis included 15,074 cases, 18,618 proxy cases, and 449,056 controls, and there were 17,410,431 genotyped and imputed SNPs tested for association with PD. Recruitment and genotyping quality control are described in the original report.^41^ Only exposure SNPs from fold 1 were used in the replication analysis, and we excluded SNPs on the X chromosome and palindromic SNPs.

### Sensitivity analyses

If tobacco use truly has a protective effect on PD, we would expect genetic variants that increase the amount of tobacco used should decrease PD risk in tobacco users, but should have no effect on PD risk in non-users. If these variants are protective for PD in separate analyses of both ever-smokers and never-smokers, however, this suggests that the protective effect is mediated by a pleiotropic pathway unrelated to tobacco use. Given that ever/never smoker status has a positive genetic correlation with “number of cigarettes smoked per day” (ρg=0.366, p=4.5×10-4), we re-ran MR analyses separately for ever-smokers and never-smokers.^42^ We also repeated this analysis using a single SNP (rs16969968) that leads to a missense mutation in *CHRNA5* and has been shown to affect the amount of tobacco use within tobacco users.^43^ In both analyses, we re-computed the effect of our instrument variables on PD in the same cohort, but stratifying on tobacco use status. We employed the same estimates of the effect of the tobacco use SNPs as in the main analysis. For the *CHRNA5* SNP, we used the estimate from Millard et al. that each allele that increases tobacco use is associated with an odds ratio of 1.21 for being a “heavy smoker” (95% CI: 1.19-1.23).^43^

### Statistical analysis

Analyses were performed using R statistical software 3.3.2 (2016-10-31).

## Results

### Tobacco use

385 SNPs were associated with self-reported smoking status (ever-versus never-smoker) in fold 1 (422 SNPs in fold 2, Tables 1 and 2 in Supplementary Data). This instrument explained an average of 1.16% of the variance in tobacco smoking liability across folds (fold 1 = 1.12%, fold 2 = 1.19%). The F statistic (F= 54.3 in fold 1, F=52.6 in fold 2) was high; validating this instrument for these analyses.

In the IVW analysis, ever-smoking caused a reduction in PD risk with an OR 0.955 (95% CI 0.921-0.991, p = 0.013) (Fig 1). There was no clear evidence of heterogeneity in estimates derived from individual SNPs (fold 1-versus-fold 2: Cochran’s Q = 184.3, p = 1.000; fold 2-versus-fold 1: Cochran’s Q = 182.4, p = 1.000). The MR-Egger intercept was not significant (p=0.193) and the estimate from MR-Egger was OR 0.867 (95% CI 0.747-1.006, p = 0.061). Funnel plots (Fig 2 A to B) suggested that individual variants were symmetrically distributed around the point estimate. Together these findings suggest no meaningful bias through unbalanced horizontal pleiotropy. The weighted median analysis was similar to the IVW estimate, suggesting minimal bias due to invalid instruments (OR = 0.927, 95% CI 0.857-1.002, p = 0.058). We replicated the IVW results in the IPDGC dataset (OR = 0.865, 95% CI 0.756-0.990, p = 0.035) and a meta-analysis of the results from the two datasets yielded a stronger p-value than results from either dataset individually (OR = 0.949, 95% CI 0.916-0.983, p = 0.003) (Fig 1).

**Figure 1:**
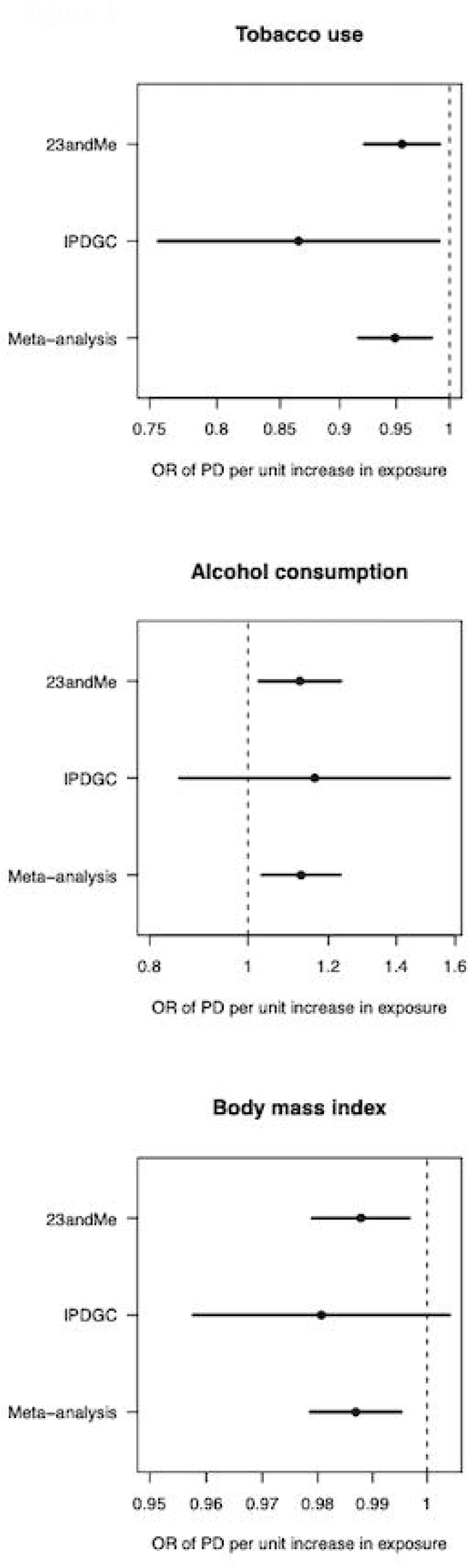
Forest plot of Mendelian randomisation causal association estimates between risk of Parkinson’s disease and unhealthy behaviours derived from meta-analysis of the 23andMe and IPDGC datasets. The pooled odds ratio (OR), derived from meta-analysis of the inverse variance weighted estimates, and 95% confidence intervals are shown. For smoking, the unit of exposure is never versus ever smoking. For alcohol, the unit of exposure is 1-group difference in daily alcohol intake. For BMI, the unit of exposure is 1 kg/m^2^. OR = odds ratio; PD = Parkinson’s disease.

**Figure 2:**
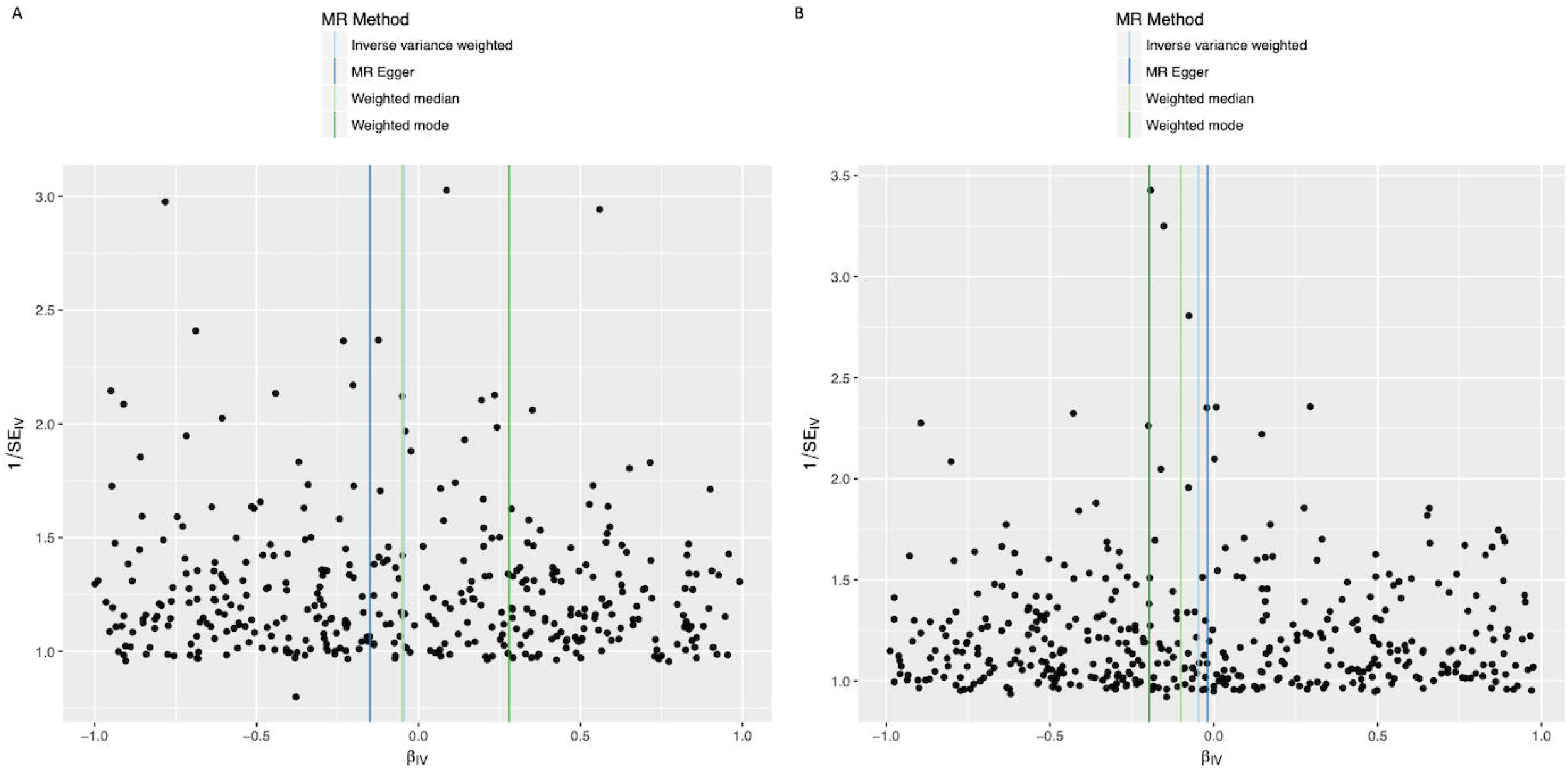
Funnel plots of individual variant effects for the smoking instrument (from fold 1 (A) and fold 2 (B)) plotted against the inverse of their standard error.

As a sensitivity analysis, we re-ran MR in the 23andMe dataset separately for ever-smokers and never-smokers. As expected, we found a similar protective effect, although less precisely estimated, in ever-smokers (OR = 0.966, 95% CI 0.885-1.055) and little evidence of a protective effect in never-smokers (OR = 1.030, 95% CI 0.966-1.097). We found similarly imprecise estimates when performing MR using a SNP in *CHRNA5* that strongly influences the amount of tobacco use within tobacco users (ever-smokers: OR = 1.037, 95% CI 0.888-1.211; never-smokers: OR = 1.033, 95% CI 0.849-1.256).

### Alcohol intake

129 SNPs were associated with self-reported alcohol in fold 1 (124 SNPs in fold 2, Tables 3 and 4 in Supplementary Data). This instrument explained an average of 0.53% of the variance in alcohol intake liability across folds (fold 1 = 0.53%, fold 2 = 0.50%). The F statistic (F=49.5 in fold 1, F=48.3 in fold 2) again validating this instrument for alcohol intake.

**Table 3.**
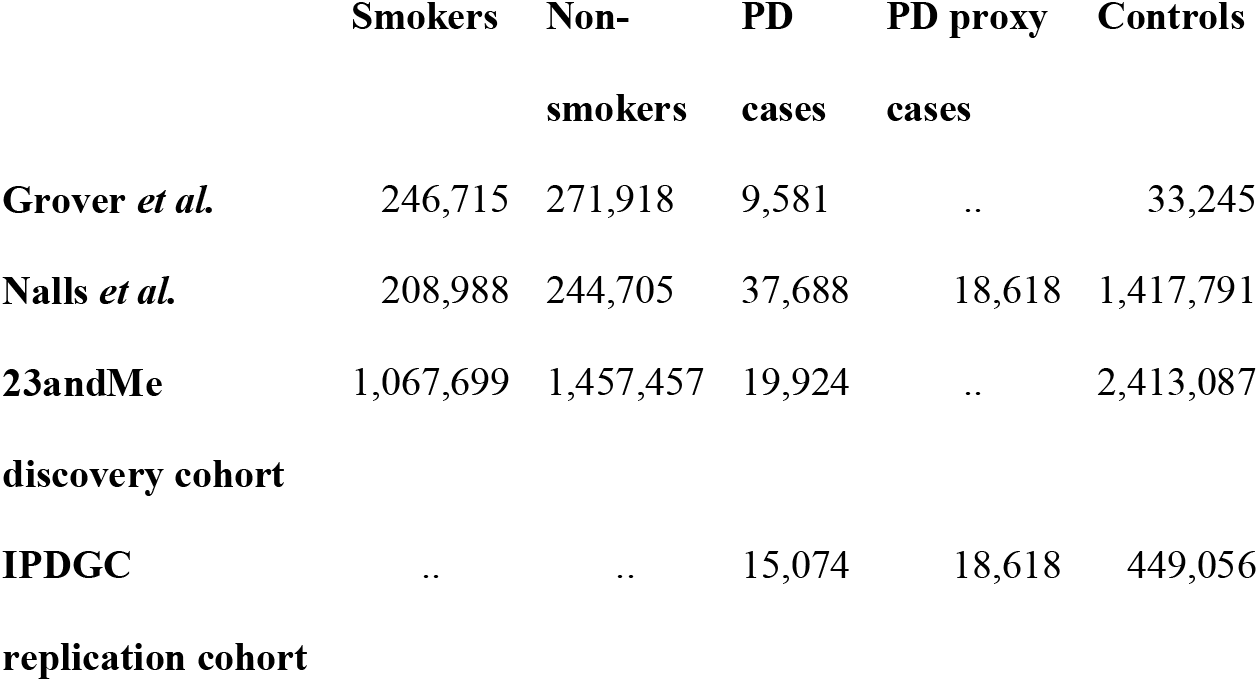
Table legend: Exposure and outcome sample sizes in the present study (23andMe discovery cohort and IPDGC replication cohort), and the two other Mendelian randomisation studies exploring the association between smoking and PD. Abbreviations: PD = Parkinson’s disease; IPDGC = International Parkinson’s disease Genomics Consortium.

In the IVW analysis, alcohol intake caused an increase in PD risk with an OR 1.125 for a 1-group increase in daily alcohol intake (95% CI 1.025-1.235, p = 0.013) (Fig 1). There was no clear evidence of heterogeneity against estimates derived from individual SNPs (fold 1-versus-fold 2: Cochran’s Q = 75.3, p = 1.000; fold 2-versus-fold 1: Cochran’s Q = 70.9, p = 1.000). The MR-Egger intercept was not significant (p=0.152) and the estimate from MR-Egger was OR 1.438 (95% CI 1.014-2.038, p = 0.041). Funnel plots (Fig 3A to B) suggested that individual variants were symmetrically distributed around the point estimate, again suggesting no meaningful bias through unbalanced horizontal pleiotropy. The weighted median estimate was consistent with the IVW estimate, but with wider confidence intervals, suggesting minimal bias due to invalid instruments (OR = 1.126, 95% CI 0.943-1.345, p = 0.189). The point estimate in the IPDGC dataset was similar to the 23andMe result, but the confidence intervals were wide (OR = 1.163, 95% CI 0.856-1.580, p = 0.334). A meta-analysis of the results from the two datasets yielded a stronger p-value than results from either dataset individually (OR = 1.128, 95% CI 1.032-1.233, p = 0.008) (Fig 1).

**Figure 3:**
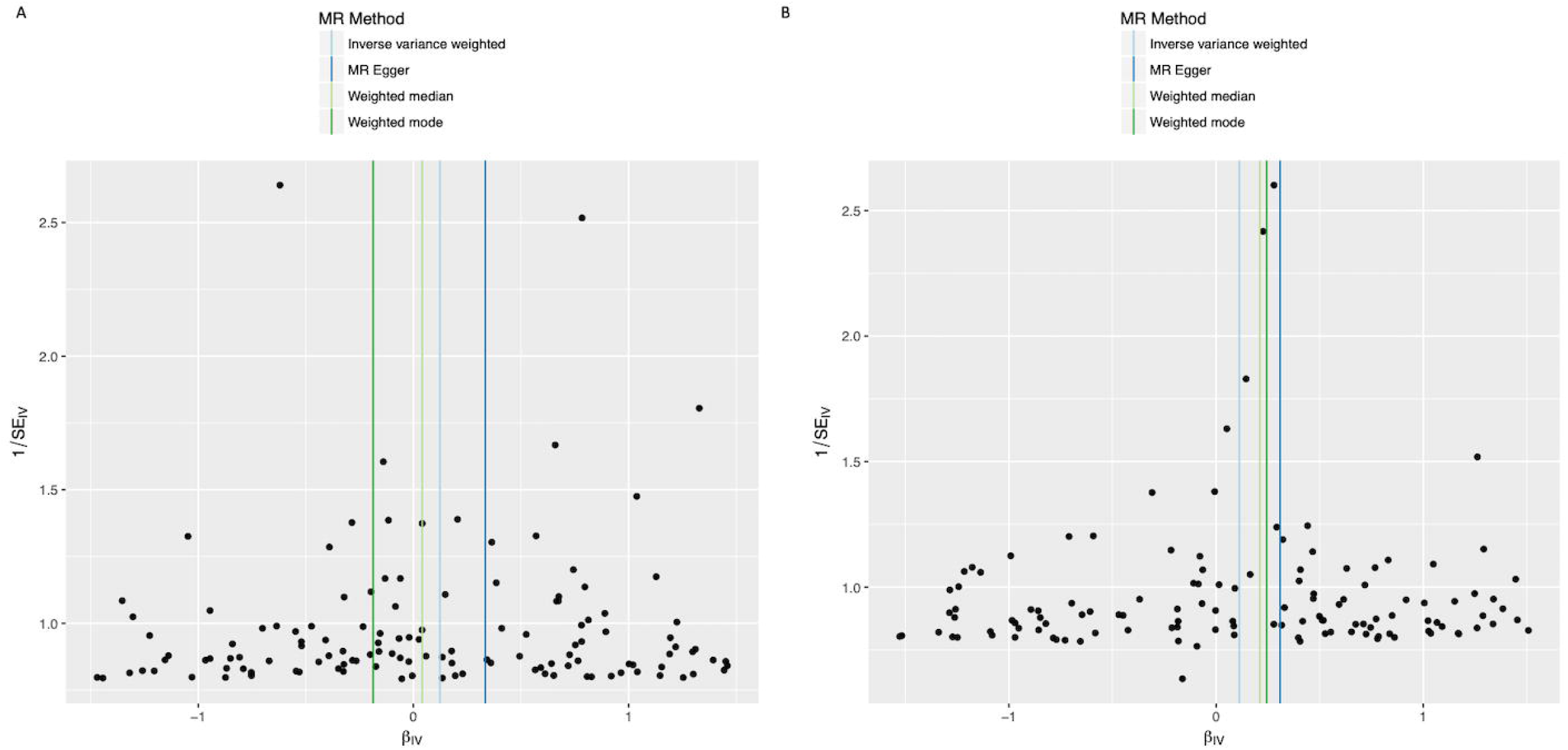
Funnel plots of individual variant effects for the alcohol instrument (from fold 1 (A) and fold 2 (B)) plotted against the inverse of their standard error.

### BMI

729 SNPs were associated with self-reported alcohol in fold 1 (693 SNPs in fold 2, Tables 5 and 6 in Supplementary Data). This instrument explained an average of 5.31% of the variance in BMI liability across folds (fold 1 = 5.31%, fold 2 = 5.15%). The F statistic (F=99.7 in fold 1, F=102.2 in fold 2) indicating the validity of this instrument for BMI.

In the IVW analysis, a genetically-estimated 1 kg/m^2^ increase in BMI causally reduced PD risk with an OR 0.988 (95% CI 0.979-0.997, p = 0.008) (Fig 1). There was no clear evidence of heterogeneity against estimates derived from individual SNPs (fold 1-versus-fold 2: Cochran’s Q = 184.3, p = 1.000; fold 2-versus-fold 1: Cochran’s Q = 539.8, p = 1.000). The MR-Egger intercept was not significant (p=0.223) and the estimate from MR-Egger was OR 0.977 (95% CI 0.957-0.997, p = 0.023). Funnel plots (Fig 4A to B) suggested that individual variants were symmetrically distributed around the point estimate, again suggesting no meaningful bias through unbalanced horizontal pleiotropy. The weighted median estimate was consistent with the IVW estimate, suggesting minimal bias due to invalid instruments (OR = 0.985, 95% CI 0.965-1.005, p = 0.132). The causal OR in the IPDGC dataset was similar to the 23andMe result, but the confidence intervals were wide (OR = 0.981, 95% CI 0.958-1.004, p = 0.106). A meta-analysis of the results from the two datasets yielded a stronger p-value than results from either dataset individually (OR = 0.987, 95% CI 0.979-0.995, p = 0.002) (Fig 1).

**Figure 4:**
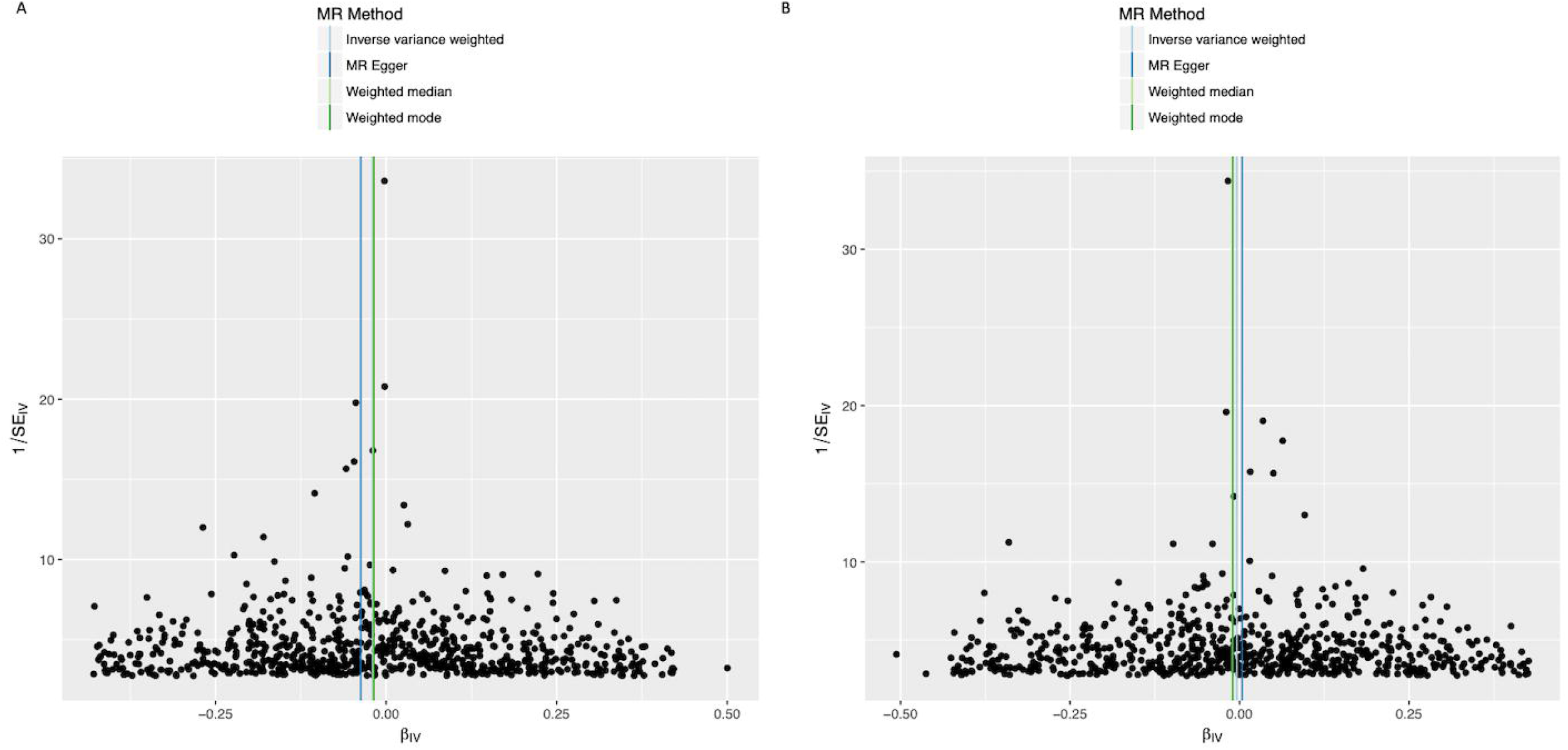
Funnel plots of individual variant effects for the BMI instrument (from fold 1 (A) and fold 2 (B)) plotted against the inverse of their standard error.

## Discussion

Using split-sample MR, we observed a protective effect of smoking on risk of PD. In a cohort of over 2.4 million participants, ever-smoking caused a 5% reduction in PD risk compared to never-smoking. Sensitivity analyses using MR-Egger and MR-median suggested that the effect was unlikely to be driven by horizontal pleiotropy, outliers, or invalid instruments. To our knowledge, this represents the largest MR study to date exploring the link between smoking and PD, and provides cautious optimism to continue to explore nicotine as a putative preventive measure.

These findings are concordant with observational studies consistently demonstrating an inverse association between smoking and PD risk.^2^ However, observational studies have not definitively ruled out reverse causality. For example, in a study of over 220,000 individuals, the protective effect of smoking on PD risk held true among ex-smokers who had quit 9-18 years before recruitment, but not among smokers who had quit >20 years before recruitment, meaning the possibility that preclinical dopaminergic changes facilitate smoking cessation could not be excluded.^44^ Similarly, a cohort study reported that parental smoking during childhood reduced future PD risk, however the possibility of a transgenerational exposure, such as a toxic agent influencing both parental smoking behaviour and PD risk in children, could not be excluded.^45^ By circumventing the use of proxy measures to overcome unmeasured confounding, residual confounding and reverse causality, the findings from MR analyses more robustly support a causal effect.

Our results are similar to those from smaller MR studies (Table 3) that have tested for a causal relationship between PD and tobacco use. Grover *et al*. found a significant protective effect (OR = 0.71 per log odds of ever smoking; 95% CI 0.57-0.90), while Nalls *et al*. found a non-significant protective effect (OR = 0.94 per log odds of ever smoking; 95% CI 0.88-1.00, p = 0.063).^41, 46^ The stringency of our instrument selection and sample size, particularly compared with the first of those studies, makes the estimate from the present study more reliable.

Early interventional studies (case series and pilot open-label trials) demonstrated beneficial effects of nicotine on motor and cognitive deficits in PD.^47-54^ However placebo-controlled studies largely failed to replicate the protective association between smoking and PD.^14-18^ This may be because trials have assessed transient changes in PD symptoms post-nicotine in small cohorts. Given the heterogeneity in PD progression, demonstrating disease modification following treatment would necessitate large sample sizes and long follow-up.^55^ An RCT currently in progress is incorporating prolonged treatment and wash-out phases pre-endpoint assessment, and will therefore better evaluate potential neuroprotective effects of nicotine on PD (ClinicalTrials.gov Identifier: NCT01560754). Trials may also have failed because they examined the disease-modifying effect of nicotine on established PD. However the mechanisms underlying PD risk may be different to those driving disease progression. Our study suggests that smoking influences the former, highlighting the need for prevention trials of candidate disease-modifying therapies in pre-manifest PD.

Mechanisms underlying the effect of smoking on PD risk remain speculative. However a disease-modifying effect of nicotine is biologically plausible given evidence that it mitigates MPTP- and 6-hydroxydopamine-induced nigrostriatal damage and motor dysfunction in rodent models and non-human primate models of PD.^56-62^

In this MR study we observed a causal increase in risk of PD with higher alcohol intake. This finding is in disagreement with a recent meta-analysis finding that alcohol consumption was associated with a lower risk of PD in retrospective case-control studies (OR for never vs. heavy/moderate drinking: 0.74; 95% CI 0.64-0.85).^63^ When the same study meta-analysed prospective cohort studies, however, the difference was not significant.^63^ It is therefore plausible that the inverse association observed in case-control studies is driven by reverse causation, whereby lower alcohol consumption in cases reflects a low dopaminergic state, suppressing addictive behaviours. Our finding is consistent with the known neurotoxic effect of excess alcohol consumption in human neuropathological studies, and with experimental studies in rodents showing that alcohol reduces dopamine in the midbrain and increases oxidative damage in nigral cells.^64, 65^ Our results suggest that alcohol consumption is unlikely to help prevent PD, and may increase risk of PD.

We have previously observed a causal protective effect of higher BMI on risk of PD, an effect that was not clearly explained by survival bias.^9^ More recently, we used a hypothesis-free approach to study causal associations between a range of exposures and PD using MR, and observed that most of the top hits related to a protective effect of increased adiposity.^66^ The results of the current study, undertaken in a large case-control settings but with some overlapping samples, support these previous observations.

Limitations of our study include that we did not explore whether effects differed between sub-groups (for example between genders, as previously suggested for the association between PD and smoking).^2^ In addition, the MR analysis assumed linearity, precluding us from identifying non-linear exposure-outcome associations. Finally, it has been argued that survival bias may distort MR analyses. However our study used a two-sample design, which has been shown to minimise the impact of survival bias on effect estimates.^67^

In conclusion, we provide evidence to support a causal protective effect of smoking and high BMI on PD risk. Conversely, we observed a causal detrimental effect of alcohol on PD risk. Although a better understanding of the underlying mechanisms, as well as development of safe delivery methods is necessary, such findings help guide the prioritisation of candidate neuroprotective approaches for RCTs in participants at higher risk of developing PD.

## Data Availability

Summary statistics for both the exposure and outcome GWASes for the relevant set of exposure SNPs are available in the supplementary data.

## Acknowledgements

We thank the research participants from 23andMe and the IPDGC who contributed to this work. The work was financially supported by The Michael J. Fox Foundation for Parkinson’s Research (grant MJFF12737), National Institute on Aging (NIA) part of the National Institutes of Health (NIH) and by 23andMe, Inc.

## Author Contributions

KH contributed to the conception and design of the study, acquired and analysed the data, and drafted the figures. MJ contributed to the conception and design of the study, and drafted the manuscript. SB acquired and analysed the data. PF, CB, MAN, AS, GDS and PC contributed to data analysis and editing the manuscript. AJN led the study group, contributed to the conception and design of the study, and edited the manuscript.

Members of the 23andMe Research Team (23andMe, Inc., 223 N Mathilda Ave, Sunnyvale, CA 94086, USA), who contributed to data acquisition, are: Michelle Agee, Stella Aslibekyan, Adam Auton, Robert K. Bell, Katarzyna Bryc, Sarah K. Clark, Sarah L. Elson, Kipper Fletez-Brant, Nicholas A. Furlotte, Pooja M. Gandhi, Barry Hicks, David A. Hinds, Karen E. Huber, Ethan M. Jewett, Yunxuan Jiang, Aaron Kleinman, Keng-Han Lin, Nadia K. Litterman, Marie K. Luff, Matthew H. McIntyre, Kimberly F. McManus, Joanna L. Mountain, Sahar V. Mozaffari, Priyanka Nandakumar, Elizabeth S. Noblin, Carrie A.M. Northover, Jared O’Connell, Aaron A. Petrakovitz, Steven J. Pitts, G. David Poznik, J. Fah Sathirapongsasuti, Janie F. Shelton, Suyash Shringarpure, Chao Tian, Joyce Y. Tung, Robert J. Tunney, Vladimir Vacic, and Xin Wang.

## Conflicts of Interest

There are no conflicts of interest to report which could bias the authors in the conduct of the reported work.

## Notes

### Competing Interest Statement

The authors have declared no competing interest.

### Funding Statement

Drs. Heilbron, Fontanillas, Cannon, and members of the 23andMe Research Team are employees of and have stock, stock options, or both, in 23andMe. Dr Noyce works at the Preventive Neurology Unit which is funded by the Barts Charity (reference MGU0364). Dr Noyce reports additional grants from Parkinson's UK, Virginia Kieley Benefaction, grants and non-financial support from GE Healthcare, and personal fees from Profile, Roche, Biogen, Bial and Britannia, outside the submitted work. Dr Nalls' participation in this work was supported in part by a consulting contract with the Intramural Research Program of the National Institute on Aging (NIA), Department of Health and Human Services; under project Z01-AG000949-02. Dr Nalls is also a founding partner at Data Tecnica International LLC, which during the time of this report has consulted for the National Institutes of Health (USA), the Michael J. Fox Foundation, Illumina Inc., Vivid Genomics, Lysosomal Therapeutics Inc., Neuron 23 Inc. and Aspen Biosciences among others, all of which had no impact on his participation in this work.

